# Attractin-Like Protein 1 (ATRNL1): An Essential Partner of MC4R to Regulate Body Weight

**DOI:** 10.64898/2026.01.12.26343722

**Authors:** Paul Buscaglia, Kate R. Bowman, Katherine Lawler, Ciria C. Hernandez, Jacopo Scotucci, Rebecca Bounds, Julia M. Keogh, Elana Henning, Roger D. Cone, I. Sadaf Farooqi, Julien A. Sebag

## Abstract

The melanocortin 4 receptor (MC4R) plays a critical role in the central control of energy homeostasis and its disruption causes severe early onset obesity. The MC4R agonist Imcivree has been approved for the treatment of certain genetic obesity syndromes. Here we demonstrate that the membrane-spanning protein Attractin-like protein 1 (ATRNL1) directly interacts with MC4R to amplify its signaling in cells and that expression of ATRNL1 potentiates the activation of MC4R neurons by MC4R agonists in mice. Disruption of ATRNL1 in the PVN of the hypothalamus causes increased food intake and obesity, which cannot be rescued by MC4R agonist treatment. In exomes from 927 children with severe early-onset obesity, we identified 21 rare ATRNL1 variants. Eleven of 17 variants studied in cells impaired ATRNL1-mediated regulation of MC4R signaling, **including variants ultra-rare or absent from UK Biobank and other populations**. Cumulatively, these findings establish ATRNL1 as an important regulator of mammalian energy homeostasis.

## Main

Central melanocortin signaling plays a critical role in the regulation of energy homeostasis^1–4^. One essential component of this machinery is the Melanocortin-4 receptor (MC4R), a Gα_s_-coupled receptor expressed throughout the hypothalamus, limbic region, and brainstem, including neurons of the paraventricular nucleus (PVN) of the hypothalamus ^5–8^. In humans, loss of function mutations in *MC4R* cause increased food intake (hyperphagia) and severe early onset obesity and are found at a prevalence of 1 in 300 in the population, 1% of people with obesity and up to 5% of children with severe early onset obesity, thus representing the most common monogenic cause of obesity ^9,10^. Gain of function MC4R variants are associated with significant protection against obesity and type 2 diabetes, as such targeting MC4R signaling has been a major focus of drug discovery efforts^11^. Recently, a MC4R agonist, Setmelanotide, was approved for the chronic weight management of children and adults with genetic disorders disrupting the MC4R pathway ^12–14^. To enable the discovery of the next generation of drugs targeting MC4R to promote weight loss, an improved understanding of the factors which regulate MC4R signaling and activity is essential.

GPCR trafficking, signaling and ligand specificity can be modulated by a range of single membrane-spanning accessory proteins. For example, the Melanocortin Receptor Accessory Protein 2 (MRAP2) acts as an endogenous potentiator of MC4R function ^15,16^. Deletion of MRAP2 causes obesity in mice and loss of function mutations are associated with obesity in humans ^17–20^. In this study we investigate the role of Attractin-like protein 1 (ATRNL1), a single transmembrane protein first identified as a binding partner of MC4R in a yeast-two-hybrid screen^21,22^, and for which the physiological function is unknown. ATRNL1 is highly homologous to Attractin, which is essential for the pigment type switching caused by agouti, a ligand of the Melanocortin 1 receptor expressed in dermal melanocytes ^23,24^. Our goal was to examine how ATRNL1 affects MC4R signaling in cells, mice and humans.

### Attractin-like 1 protein potentiates MC4R signaling

The interaction of MC4R and ATRNL1 was first identified through a yeast two-hybrid screen for MC4R interacting proteins^21^. We first verified that MC4R and ATRNL1 can form a complex when expressed in mammalian cells by transfecting CHO cells with constructs encoding N-terminally tagged human MC4R (3HA-hMC4R) and human ATRNL1 (hATRNL1-3XFlag) or empty vector. Cells were lysed and MC4R or ATRNL1 were pulled down from lysates using anti-HA or anti-Flag antibody respectively before detection of ATRNL1 by western blot. ATRNL1 was detectable both in the Flag and HA pull downs (Fig. 1A), demonstrating that ATRNL1 and MC4R form a complex. To determine if this complex exists in live cells, we used the Nanobit luciferase recombination assay, which relies on the rapid recombination of two fragments, LgBiT and SmlBiT, of nanoluciferase fused to proteins of interest. Cells were transfected with either MC4R-LgBiT alone or MC4R-LgBiT and ATRNL1-SmlBiT. Upon addition of substrate, luminescence was readily detectable in live cells transfected with both fusion proteins (Fig. 1B), confirming the formation of the MC4R/ATRNL1 complex in live cells.

**Figure 1:**
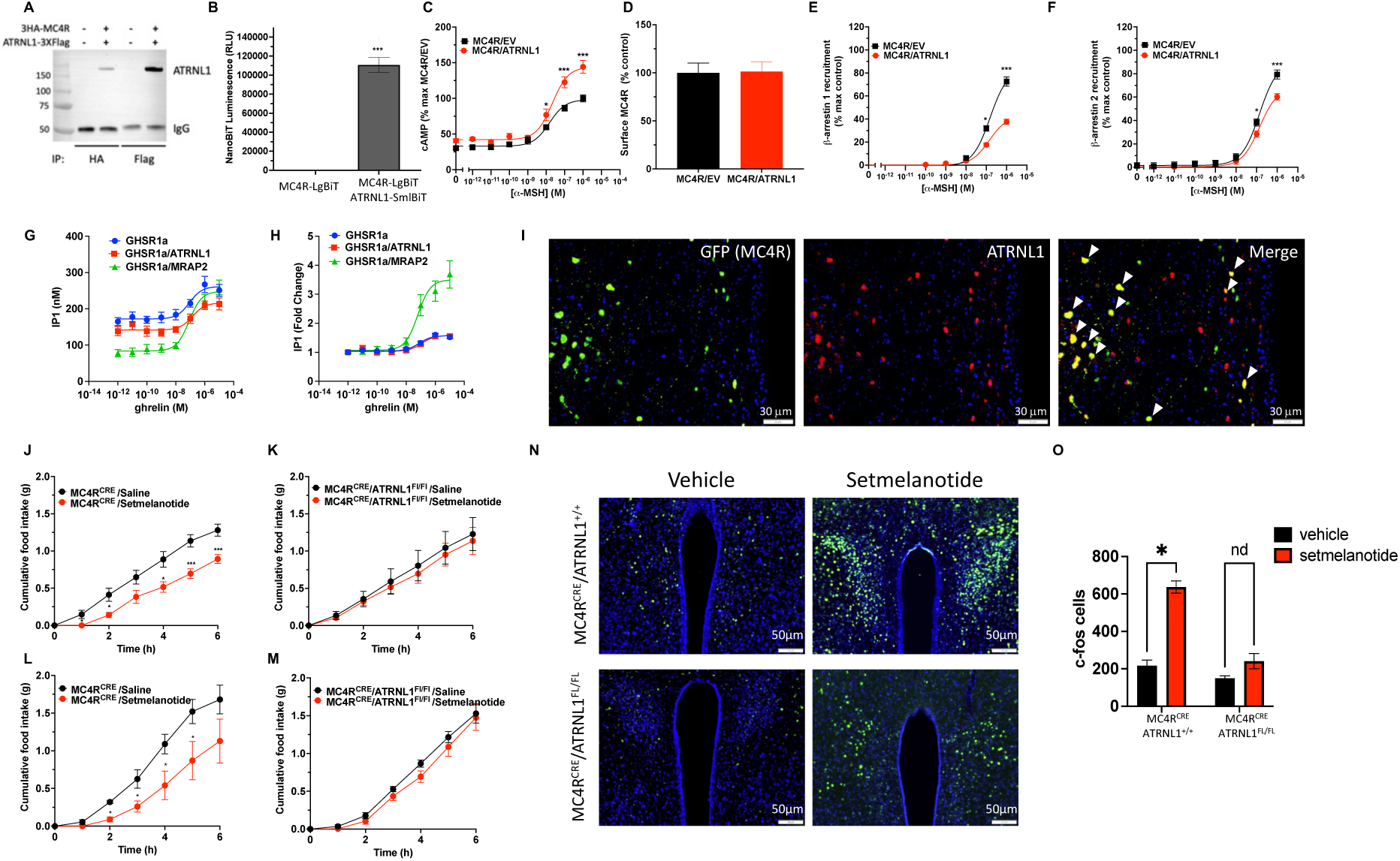
ATRNL1 enhances MC4R signaling. **A.** Western Blot detection of ATRNL1-3xFlag from lysates of HEK293 cells transfected with empty vector or with 3HA-MC4R and ATRNL1-3xFlag following immunoprecipitation with either anti-HA (MC4R) or anti-Flag (ATRNL1). **B.** NanoBit Assay in HEK293 cells transfected with MC4R-LrgBiT alone or with ATRNL1-SmlBiT(N=3; n=3-5 per group). **C.** a-MSH-stimulated cAMP assay in HEK293 cells transfected with MC4R and either empty vector (EV) or ATRNL1 using the GloSensor luminescent assay. (N=3; n=5 per group). **D.** Surface expression of HiBiT-MC4R in the presence or absence of ATRNL1 in HEK293 cells N=3; n=5 per group) **E-F**. a-MSH-stimulated b-arrestin1 (**E**) or b-arrestin2 (**F)** recruitment to MC4R in the presence or absence of ATRNL1. (N=3; n=4-5 per group). **G-H.** IP1 accumulation measured in CHO cells transfected with GHSR1a alone or with either MRAP2 or ATRNL1 and treated with inducated ghrelin concentration. Data are shown as IP1 concentration (**G**) or fold change (**H**) to illustrate the constitutive activity or efficacy changes respectively. **I.** *In Situ* hybridization detection of ATRNL1 in brain slices of MC4RCRE mice injected with AAV1-CMV-Flex-eGFP in the PVN. (n=3 per group). **J-M**. Cumulative food intake measured up to 6 hours in MC4RCRE/ATRNL1+/+ **(J)** and MC4RCRE/ATRNL1Fl/Fl **(K)** male mice (n=5 per group) and in MC4RCRE/ATRNL1+/+ **(L)** and MC4RCRE/ATRNL1Fl/Fl **(M)** female mice (n=4-9 per group) injected IP with saline or 2 mg/kg Setmelanotide. **N.** cFos detection in brain slices of MC4RCRE/ATRNL1+/+ and MC4RCRE/ATRNL1Fl/Fl mice injected IP with saline or 2 mg/kg Setmelanotide. (n=6 per group). **O**. Quantitation of PVN cFos positive cells in MC4RCRE/ATRNL1+/+ and MC4RCRE/ATRNL1Fl/Fl injected with vehicle or Setmelanotide (n=6 per group). Mann-Whitney test, Unpaired and Non-Parametric*p<0.05 **p<0.01***p<0.001

To test whether ATRNL1 alters MC4R signaling, we transfected CHO cells with the GloSensor luminescent cyclic AMP (cAMP) reporter, MC4R and either empty vector or ATRNL1. In cells transfected with MC4R alone, the agonist α-MSH increased intracellular cAMP production in a concentration-dependent manner. Co-transfection with ATRNL1 significantly increased the efficacy but did not significantly alter the potency of α-MSH (Fig. 1C), demonstrating that while ATRNL1 is not required for MC4R signaling in cells, the presence of ATRNL1 significantly enhances MC4R signaling through Gα_s_. We observed no difference in surface MC4R density between control and ATRNL1 expressing cells (Fig. 1D), suggesting that the enhanced signaling measured in ATRNL1 containing cells is not due to increased receptor number at the plasma membrane.

β-arrestins are intracellular proteins which mediate desensitization, internalization and signaling by GPCRs including MC4R. To assess the possible role of ATRNL1 in regulating β-arrestin recruitment to MC4R, we used the Nanobit β-arrestin recruitment assay. CHO cells transfected with MC4R-LgBiT, β-arrestin1 or β-arrestin2 N-terminally fused to SmlBiT, and either ATRNL1 or empty vector, were treated with increasing concentrations of αMSH. We found that the presence of ATRNL1 resulted in a significant reduction of both β-arrestin 1 (Fig. 1E) and β-arrestin 2 (Fig. 1F) recruitment to the receptor with a more pronounced inhibition of β-arrestin1 recruitment compared to

β-arrestin2 (40% and 20% reduction respectively). The inhibition of β-arrestins recruitment to MC4R by ATRNL1 would be expected to reduce receptor desensitization and thereby may explain the amplification of cAMP production downstream of MC4R activation.

To determine if, like MRAP2, ATRNL1 is a promiscuous GPCR regulator, we tested the effect of ATRNL1 on signaling downstream of the ghrelin receptor (GHSR1a). We have previously shown that MRAP2, in addition to its potentiating effect on MC4R signaling^15^, drastically alters GHSR1a signaling by reducing its constitutive activity and enhancing its response to ghrelin^25–28^. In contrast to MRAP2, ATRNL1 expression did not significantly alter the constitutive activity (Fig. 1G) or efficacy (Fig. 1H) of ghrelin at GHSR1a. These results suggest that ATRNL1 may be less promiscuous than MRAP2, however, more GPCRs will need to be tested to determine the selectivity of ATRNL1.

### ATRNL1 is required for normal MC4R responsiveness in vivo

We next examined whether ATRNL1 is expressed in MC4R neurons and regulates their activation by MC4R agonists in mice. First, we used in-situ hybridization to detect ATRNL1 in brain slices of MC4R^CRE^ mice stereotaxically injected with AAV-FLEX-GFP in the PVN. We found that most PVN MC4R neurons, labeled with GFP, express ATRNL1 (Fig. 1I). ATRNL1 expression was however, not restricted to MC4R neurons in the PVN, suggesting that ATRNL1 may play roles other than MC4R regulation. To test the functional consequence of deleting ATRNL1 in MC4R neurons, we bred MC4R^CRE^ with ATRNL1^flox^ mice to generate MC4R^CRE^/ATRNL1^+/+^ and MC4R^CRE^/ATRNL1^Fl/Fl^ animals. Male and female mice of each genotype were fasted overnight and injected with vehicle or the MC4R agonist setmelanotide before returning food to the cage and measuring food intake. Whereas setmelanotide significantly reduced food intake in both male and female control animals, deletion of ATRNL1 in MC4R neurons abrogated the anorexigenic effect of the MC4R agonist (Fig. 1J-M), demonstrating that ATRNL1 is required for MC4R-mediated inhibition of food intake. To further investigate the requirement of ATRNL1 for MC4R neurons activation, fasted MC4R^CRE^/ATRNL1^+/+^ and MC4R^CRE^/ATRNL1^FL/FL^ mice were injected with either saline or the Setmelanotide before staining brain slices for cFos and counting the number of activated neurons in the PVN. As expected, administration of the MC4R agonist setmelanotide resulted in activation of MC4R neurons in the PVN of control MC4R^CRE^/ATRNL1^+/+^ mice (Fig. 1N and O). This response was however largely abrogated in MC4R^CRE^/ATRNL1^Fl/Fl^ animals (Fig. 1N and O), further demonstrating the importance of ATRNL1 for MC4R function in vivo.

To assess the physiological impact of deleting ATRNL1 in the PVN of adult mice, we stereotaxically injecting AAV coding for either GFP (control) or CRE recombinase bilaterally in male and female ATRNL1^Fl/FL^ animals (Fig. 2A and B). Two weeks after virus injection, both male and female ATRNL1 KO^PVN^ mice displayed significantly increased body weight (Fig. 2C and E) and food intake (Fig. 2D and F) compared to control animals. To determine whether ATRNL1 plays an important role in the anorexigenic response to MC4R stimulation, male and female ATRNL1^Fl/FL^ mice were injected with AAV-GFP or AAV-CRE bilaterally in the PVN and cannulated in the lateral ventricle. Two weeks after surgery, animals were single housed in cages containing a FED3.1 automated food intake monitoring system. After habituation, mice were administered either the MC4R agonist Melanotan II (MTII) or saline (control) via intracerebroventricular injection (ICV), one hour before the start of the dark phase. As expected, MTII caused a significant decrease in food intake in both male and female control mice (Fig. 2G and I). In contrast, the anorexigenic effect of MTII was significantly reduced in animals lacking ATRNL1 in the PVN (Fig. 2H and J). This result is consistent with the in vitro data showing that ATRNL1 is important for MC4R signaling and further establishes the importance of ATRNL1 for the anorexigenic activity of MC4R in vivo.

**Figure 2:**
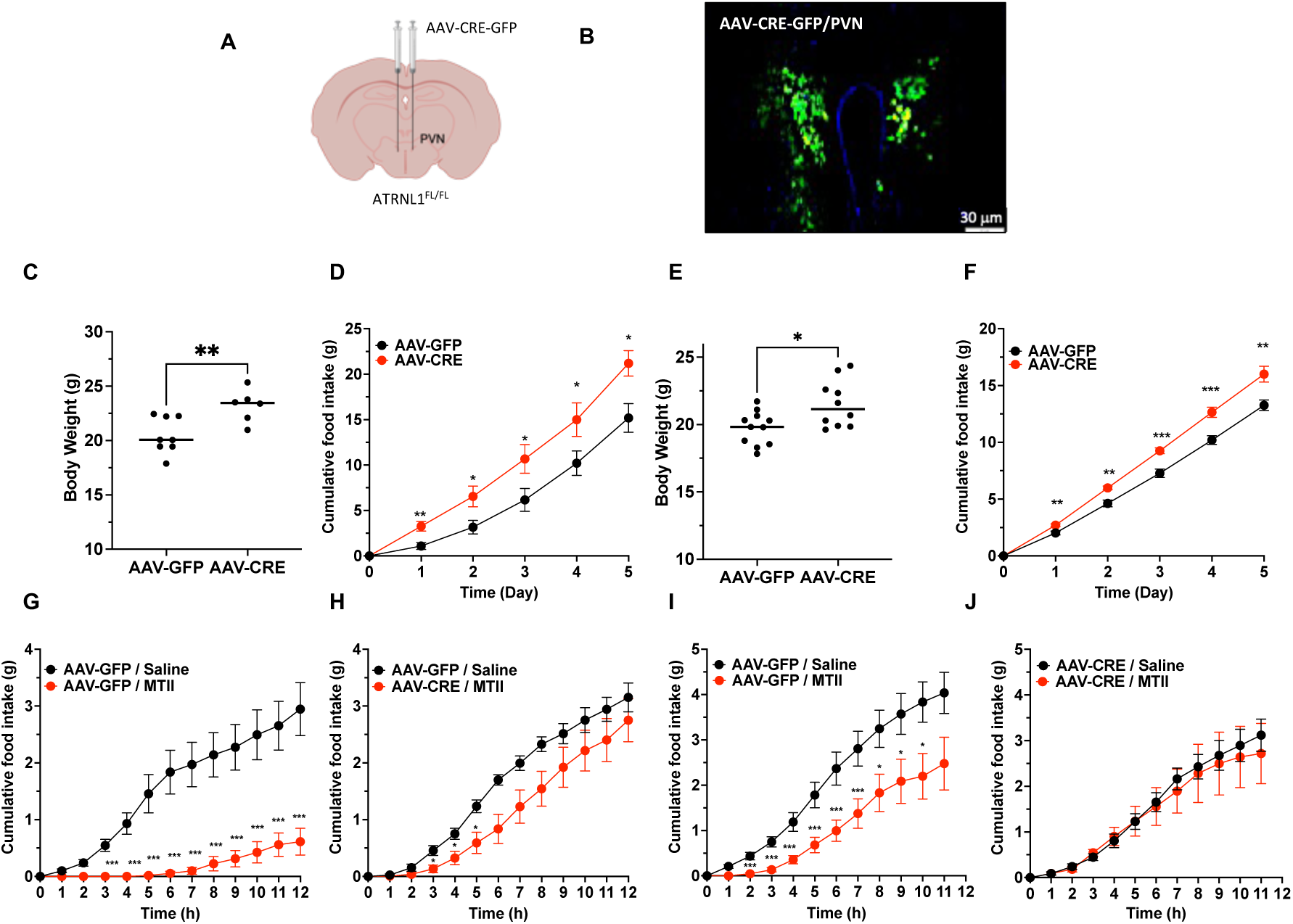
Deletion of ATRNL1 in PVN promotes hyperphagia, weight gain and impairs MC4R function. **A**. Schematic representation of bilateral PVN injection of AAV1-GFP or AAV1-Cre-GFP. **B**. Validation of stereotaxic targeting in the PVN using GFP fluorescence. **C and E**. Body weight of males (**C**) and females (**E**) ATRLN1^Fl/Fl^ mice bilaterally injected with AAV1 coding for either GFP (control) or CRE in the PVN two weeks post injection n=6-8 per group. **D and F.** Cumulative food intake in males (**D**) and females (**F**) ATRNL1^Fl/Fl^ mice injected with AAV1-GFP or AAV1-CRE-GFP bilaterally in the PVN. n=9-10 per group. **G-H.** Cumulative food intake in male ATRNL1^Fl/Fl^ mice injected with AAV1-GFP (**G**) or AAV1-CRE-GFP (**H)** bilaterally in the PVN following ICV injection of saline or 4 mg MTII at the beginning of the dark phase. n=12-14 per group. **I-J.** Cumulative food intake in female ATRNL1^Fl/Fl^ mice injected with AAV1-GFP (**I**) or AAV1-CRE-GFP (**J)** bilaterally in the PVN following ICV injection of saline or 4 mg MTII at the beginning of the dark phase. n=7-10 per group. Mann-Whitney test, Unpaired and Non-Parametric *p<0.05 **p<0.01 ***p<0.001

To directly test the effects of disrupting ATRNL in MC4R expressing neurons, we bred MC4R^CRE^ mice with ATRNL1^flox^ mice. Similar to animals with PVN targeted deletion of ATRNL1, MC4R^CRE^/ATRNL1^Fl/Fl^ mice displayed a significant increase in food intake (Fig. 3A and D). This increased feeding was further exacerbated when animals were fed a high fat diet (HFD) (Fig. 3B and E). Both male and female MC4R^CRE^/ATRNL1^Fl/Fl^ mice weighed more than control MC4R^CRE^ animals from a young age (4 weeks old) and maintained an elevated weight (Fig. 3C and F). Upon switching to a high fat diet, both control and MC4R^CRE^/ATRNL1^Fl/Fl^ gained weight at an increased rate, however, the weight gain was exaggerated in MC4R^CRE^/ATRNL1^Fl/Fl^ animals (Fig. 3C and F). Since MC4R is also known to regulate energy expenditure, male and female MC4R^CRE^ and MC4R^CRE^/ATRNL1^Fl/Fl^ mice were placed in Sable Promethion metabolic cages to measure energy expenditure. We found no difference in oxygen consumption between genotypes suggesting that ATRNL1 does not significantly alter energy expenditure (Fig. 3G and H). We also found no significant difference in glucose tolerance in male or female MC4R^CRE^ and MC4R^CRE^/ATRNL1^Fl/Fl^ mice, regardless of diet (Fig. 3I-L). To verify that deletion of ATRNL1 in MC4R neurons specifically impaired MC4R signaling rather than cause neuronal damage, we tested whether an anorexigenic response in those mice could still be achieved by activating MC4R neurons through a different mechanism. For this, MC4R^CRE^/ATRNL1^Fl/Fl^ mice were stereotaxically injected with AAV-FLEX-hM3q in the PVN to express the activating DREADD in MC4R neurons. Whereas setmelanotide injection did not decrease refeeding after an overnight fast (Fig. 3M), stimulation of MC4R neurons by CNO injection significantly reduced food intake (Fig. 3N), thus confirming that neuronal function is retained but MC4R activity is impaired.

**Figure 3:**
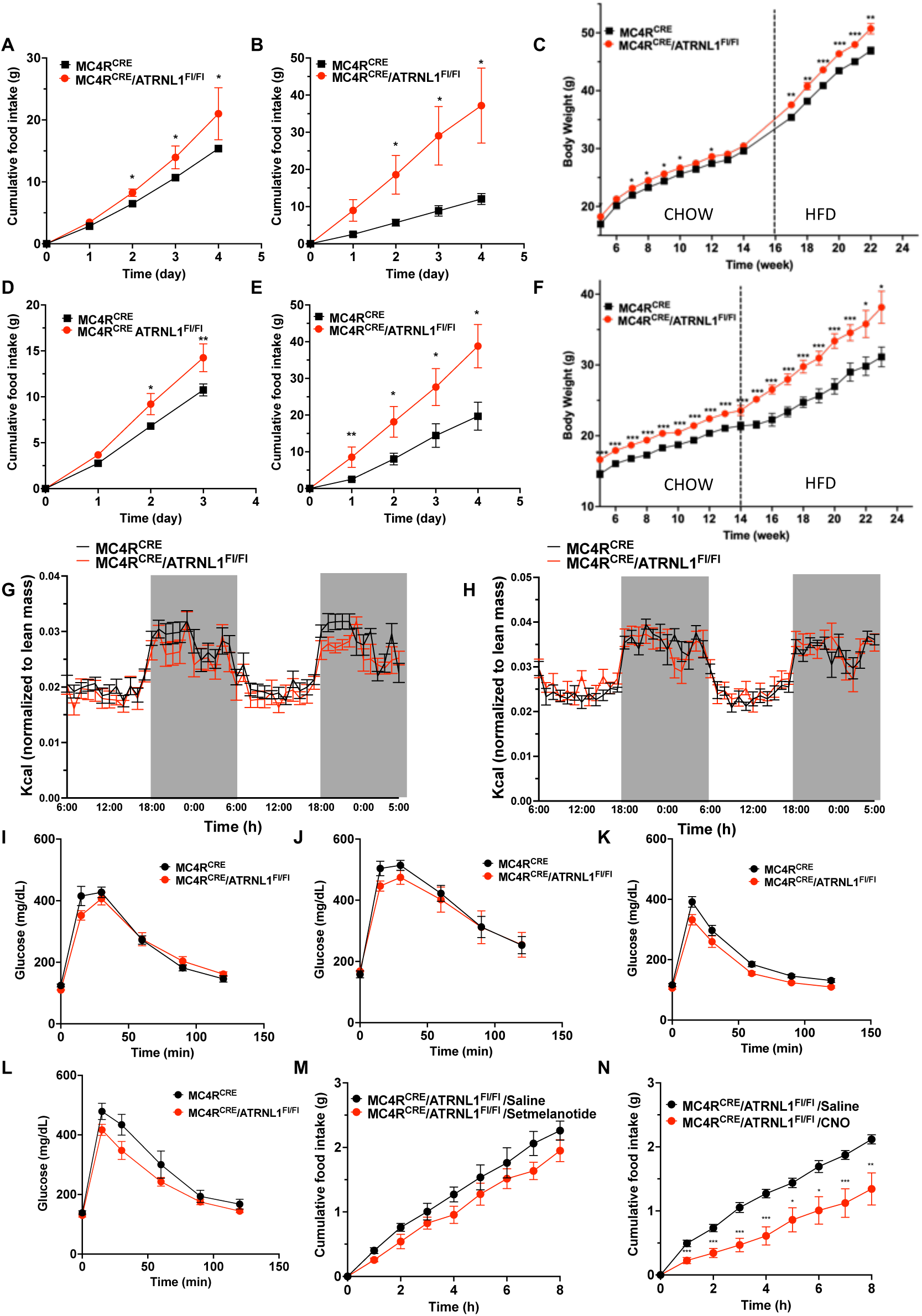
Deletion of ATRNL1 in MC4R neurons results in hyperphagia and increased body weight. **A and B.** Cumulative food intake of males MC4R^CRE^ and MC4R^CRE^/ATRNL1^Fl/Fl^ mice on standard CHOW **(A)**, n=15-17 per group and on HFD **(B)** n=9-10 per group. **C**. Body weight of male MC4R^CRE^ and MC4R^CRE^/ATRNL1^Fl/Fl^ mice on standard CHOW and HFD (dashed line indicate switch to HFD), n=12-19 per group. **D and E.** Cumulative food intake of females MC4R^CRE^ and MC4R^CRE^/ATRNL1^Fl/Fl^ mice on standard CHOW **(D)**, n=7-9 per group and on HFD **(E)** n=7-8 per group. **F**. Body weight of female MC4R^CRE^ and MC4R^CRE^/ATRNL1^Fl/Fl^ mice on standard CHOW and HFD (dashed line indicate switch to HFD), n=12-17 per group. **G and H.** Energy expenditure of MC4R^CRE^/ATRNL1^−/−^ and MC4R^CRE^/ATRNL1^Fl/F^ mice in male (G) and female mice (H) n= 6 per group. **I and J** IPGTT in male MC4R^CRE^ and MC4R^CRE^/ATRNL1^Fl/Fl^ mice fed a standard CHOW **(I)** or HFD **(J)** n=4-6 per group. **K and L.** IPGTT in female MC4R^CRE^ and MC4R^CRE^/ATRNL1^Fl/Fl^ mice fed a standard CHOW **(K)** or HFD **(L)** n=8-10 per group. **M and N.** Cumulative food intake in males MC4R^CRE^/ATRNL1^Fl/Fl^ mice on standard chow injected IP with 2mg/kg Setmelanotide or Saline **(M)** and IP 1 mg/kg CNO or Saline **(N)** n=4-8 per group following two weeks DREADD expression in the PVN. Mann-Whitney test, Unpaired and Non-Parametric. *p<0.05 **p<0.01***p<0.001

### Rare ATRNL1 variants found in people with severe obesity impair MC4R signaling

To investigate the possible role of ATRNL1 in human energy homeostasis, we interrogated whole exome sequencing data from 1,623 children with severe early-onset obesity (defined as Body Mass Index (BMI) standard deviation score > 3, onset before age 10 years) of diverse reported ancestries recruited to the Genetics of Obesity Study (GOOS). These 1,623 GOOS exomes comprised N=927 exomes from white British GOOS participants joint-called with 4,057 ancestry-matched controls (internal controls^29^) and an additional N=696 exomes from participants of white British or diverse reported ancestries. We identified 18 rare variants (allele frequency <0.1% in all gnomAD v2.1.1 exome subpopulations) in 20 unrelated GOOS participants (one individual carried two rare variants) (Table 1); 5 of these variants were also present among our white British internal control dataset (Table 1). Most mutations affected residues within the large N-terminal region of the protein; two were located in the C-terminal tail. ATRNL1 variant carriers had a history of hyperphagia and weight gain since childhood but no history of developmental delay or any other medical condition. Among the ATRNL1 variants found in GOOS, D1229N and ΔC1258-were ultra-rare or absent from UK Biobank. There was only one UK Biobank carrier of D1229N (BMI>40 kg/m^2^, severe obesity).

**Table 1:** Rare variants in ATRNL1 found in the GOOS cohort. # variant also found in controls; * two variants carried by the same individual in GOOS and by a single control (likely rare haplotype)

| Proband ID | Protein position | Residue | SIFT | PolyPhen | CADD PHRED | gnomAD_exomes_NFE_AF | gnomAD_exomes_POPMAX_AF |
| --- | --- | --- | --- | --- | --- | --- | --- |
| 1 | 103 | S/F | Deleterious low confidence (0) | Probably damaging (0.997) | 28.9 | 1.799860e-05 | 1.799860e-05 |
| 2 | 186 | Y/C | Deleterious (0) | Probably damaging (0.947) | 25.6 | 9.479930e-06 | 3.407390e-05 |
| 3 | 380 | N/S | Deleterious low confidence (0.02) | Probably damaging (0.993) | 25.4 | - | - |
| 4 | 419 | M/V | Tolerated low confidence (1) | Benign (0) | 13.53 | 7.965590e-05 | 1.964510e-04 |
| 5 | 502 | Y/H# | Deleterious low confidence (0.01) | Benign (0.222) | 27.5 | 9.626310e-06 | 6.807350e-05 |
| 6 | 574 | N/S | Tolerated low confidence (0.07) | Benign (0.013) | 19.15 | 8.879730e-06 | 8.879730e-06 |
| 7 | 613 | P/L# | Deleterious (0.04) | Possibly damaging (0.516) | 27.4 | 3.434010e-04 | 3.434010e-04 |
| 8 | 775 | Y/H | Deleterious low confidence (0) | Probably damaging(0.994) | 24.7 | 0.000000e+00 | 2.914940e-05 |
| 9 | 885 | P/L | Deleterious low confidence (0) | Possibly damaging(0.465) | 24.9 | 7.189720e-05 | 7.189720e-05 |
| 10 | 940 | P/S | Tolerated (0.16) | Benign (0.006) | 22.5 | 2.67399e-05 | 0.000133378 |
| 11 | 987 | H/R | Tolerated low confidence (0.38) | Benign (0.003) | 14.58 | 1.759480e-05 | 6.153090e-05 |
| 12 | 987 | H/R | Tolerated low confidence (0.38) | Benign (0.003) | 14.58 | 1.759480e-05 | 6.153090e-05 |
| 13 | 1154 | F/V | Deleterious (0.04) | Benign (0.214) | 23.7 | - | - |
| 14* | 1191 | Y/N# | Tolerated (1) | Benign (0.014) | 20.7 | 4.373210e-05 | 4.373210e-05 |
| 14* | 1193 | K/E# | Tolerated (0.12) | Benign (0.411) | 23.3 | 4.381070e-05 | 4.381070e-05 |
| 15 | 1198 | S/N# | Tolerated (0.98) | Benign (0.196) | 19.14 | 3.355920e-04 | 3.355920e-04 |
| 16 | 1198 | S/N | Tolerated (0.98) | Benign (0.196) | 19.14 | 3.355920e-04 | 3.355920e-04 |
| 17 | 1229 | D/N | Deleterious low confidence (0) | Benign (0.251) | 23.7 | - | - |
| 18 | 1257-1258 | TC/T | - | - | - | - | - |
| 19 | 1263 | R/Q | Deleterious low confidence (0.01) | Probably damaging (0.968) | 28 | 1.838100e-04 | 1.838100e-04 |
| 20 | 1263 | R/Q | Deleterious (0.01) | Benign (0.342) | 27.0 | 1.838100e-04 | 1.838100e-04 |

Among unrelated white British participants in UK Biobank, for each GOOS variant we could not reject the null hypothesis that mean BMI among UK Biobank carriers did not differ from non-carriers (adjusted p>0.05 for each variant; N=1 to 466 carriers per variant). There were no *ATRNL1* predicted truncating variants among GOOS exomes. In UK Biobank 500K genomes, there were 278 carriers of *ATRNL1* predicted protein-truncating variants among unrelated white British participants including rare recurrent stopgains, frameshifts and splice donor/acceptor variants (allele frequencies up to 0.003%, ENST00000355044 p.Leu546Ter, gnomAD v2.1.1 Non-Finnish European). Mean BMI among unrelated white British carriers of *ATRNL1* predicted protein-truncating variants did not differ from non-carriers in UK Biobank (p=0.5, b=0.2 [95%CI (-0.4, 0.9)], N=205 stopgain/frameshift carriers, N=335,197 non-carriers).

To test whether specific rare variants might affect the function of ATRNL1, we generated 17 mutants by targeted mutagenesis and verified their expression in transfected cells by western blot (Fig. 4A). Since MC4R stimulation results both in cAMP production and closing of the Kir7.1 potassium channel ^30–33^, we measured both αMSH-stimulated cAMP production and inhibition of potassium currents. For cAMP measurements, we used HEK293 cells stably expressing MC4R and the Glosensor 20F cAMP reporter^34^. Luminescence signal was recorded using an FDSS microcell kinetic plate reader. For the Kir7.1 assay, we used HEK293 cells stably expressing MC4R and Kir7.1^35^. Potassium currents recordings were performed using a SynchroPatch automated patch clamp system. In both cases, cells were transfected with empty vector (EV), WT ATRNL1 or ATRNL1 mutants. Whereas ATRNL1 enhanced αMSH-stimulated cAMP production downstream of MC4R, it did not alter the ability of αMSH to reduce Kir7.1 conductance (Fig 4B). Compared to WT ATRNL1, 5 ATRNL1 mutations (S103F, P885L, H987R, Y1191N and R1263Q) abolished the potentiating effect of ATRNL1 on αMSH-stimulated cAMP production, 6 mutations (N380S, Y502H, P613L, Y775H, K1198N and D1229N) displayed partial loss of function and 6 mutations (M419V, N574S, P940S, F1154V, P1193E and ΔC1258) displayed normal activity (Fig. 4B, Fig. S1). When looking at Kir7.1 conductance, we found 8 mutations that severely impaired α-MSH-mediated inhibition of potassium currents (S103F, N380S, Y502H, N574S, P613L, P885L, F1154V and ΔC1258) (Fig. 4B, Fig. S2). Neither WT ATRNL1 nor any mutant altered the density of MC4R at the cell surface as measured using N-terminally tagged HiBiT-MC4R transfected in HEK293 cells with EV, ATRNL1 or mutants ATRNL1 (Fig. 4C). Interestingly, whereas some mutation negatively impacted MC4R signaling through both pathways, like S103F (Fig. 4D and E) or P885L (Fig. 4F and G), some mutants like H987R only impaired the cAMP signaling pathway (Fig. 4H and I) while others, like ΔC1258, selectively prevented MC4R-mediated Kir7.1 closing (Fig. 4J and K). Findings were summarized in Fig. 4L. Phenotypes of carriers with ATRNL1 mutations impairing MC4R signaling through cAMP or Kir7.1 are listed in table 2.

**Figure 4:**
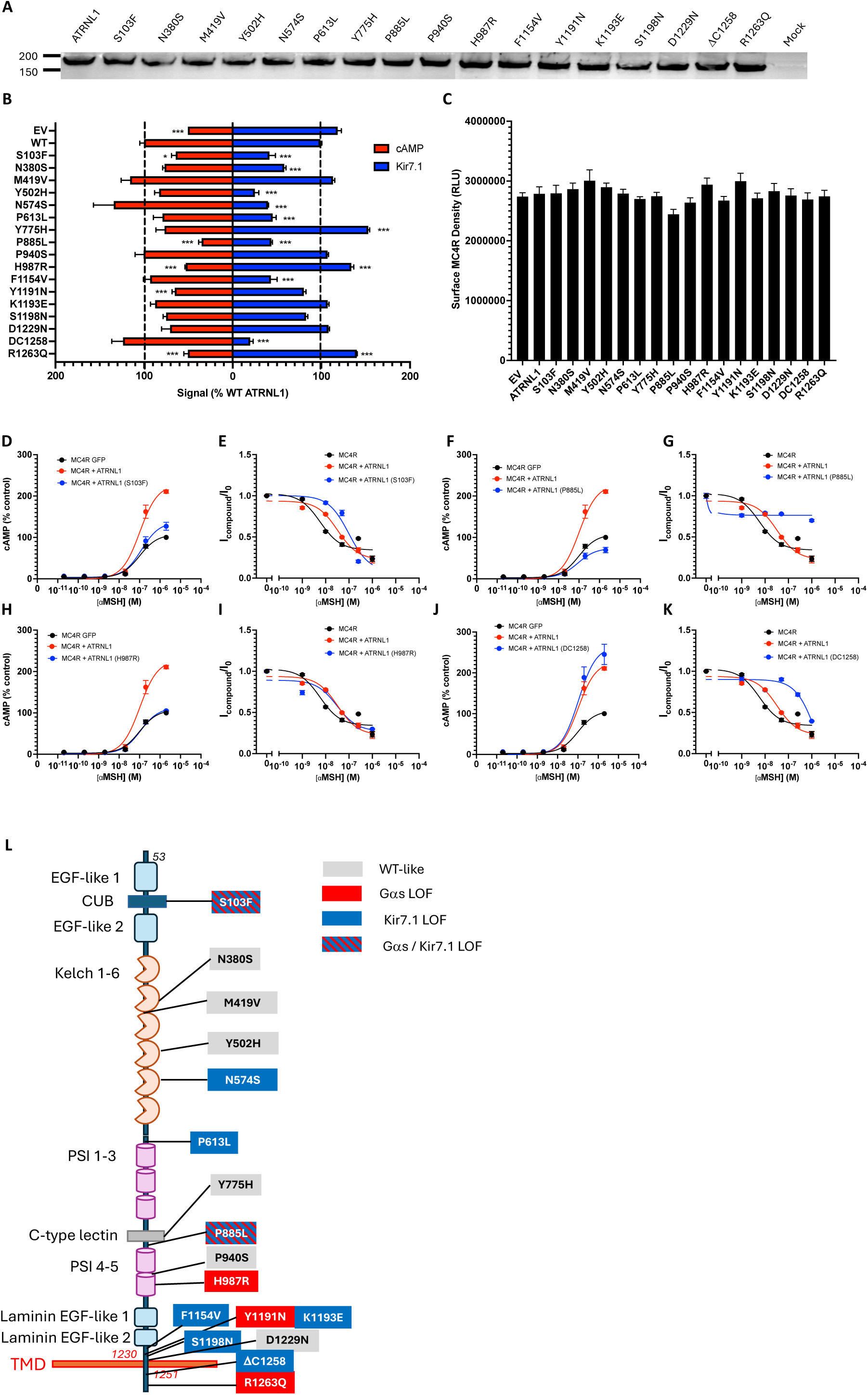
Functional impact of ATRNL1 mutations identified in human carriers. **A.** Western blot detection of WT and mutants ATRNL1 transfected in HEK-293T cells. **B**. αMSH-mediated cAMP and Kir7.1 signaling in cells expressing MC4R alone or with WT or mutants ATRNL1. cAMP was measured using GloSensor and potassium currents using Syncropatch. Data are shown in % of WT ATRNL1 control. For cAMP, the pick of the concentration response curve was used. For Kir7.1, the % inhibition was calculated using the area under the curve of the concentration response curve. **C.** Surface expression of HiBiT-MC4R expressed with ATRNL1 WT or mutant ATRNL1 in HEK293 cells using extracellular HiBiT assay. **D, F, H and J**. Representative a-MSH-stimulated cAMP production in HEK293 cell stably expressing MC4R and the GloSensor reporter, and transiently transfected with GFP (control), ATRNL1-3xFlag or the indicated ATRNL1 mutant, ATRNL1-3xFlag S103F **(D)**, ATRNL1-3xFlag P885L **(F)**, ATRNL1-3xFlag H987R **(H)**, ATRNL1-3xFlag DC1258 **(J)**. **E, G, I and K.** Representative whole-cell patch-clamp recordings showing the characteristic inwardly rectifying I–V relationship of Kir7.1 in HEK293 cells stably expressing MC4R/Kir7.1 and transiently transfected with GFP (control), ATRNL1-3×FLAG, or the indicated ATRNL1 mutant, ATRNL1-3xFlag S103F **(E)**, ATRNL1-3xFlag P885L **(G)**, ATRNL1-3xFlag H987R **(I)**, ATRNL1-3xFlag DC1258 **(K) L.** Schematic displaying ATRNL1 mutations identified in humans and their consequence on MC4R signaling. Mann-Whitney test, Unpaired and Non-Parametric. *p<0.05 ***p<0.001

**Table 2:** Phenotypes of probands in GOOS carrying loss of function ALP-1 variants. NA-not available; normal range for insulin 0-60 pmol/L

| Subject ID | Variant | cAMP | Kir7.1 | Sex | height centile | Height SDS | BMI (kg/m<sup>2</sup>) | BMI SDS | Glucose (mmol/l) | Insulin (pmol/L) | Leptin (ng/ml) |
| --- | --- | --- | --- | --- | --- | --- | --- | --- | --- | --- | --- |
| 1 | S103F | LOF | LOF | Female | 99.6 | 2.68 | 35.1 | 3.4 | 5.4 | 94 | 73.4 |
| 3 | N380S | moderate LOF | LOF | Female | 91 | 1.22 | 27 | 4.88 | 4.4 | NA | 33 |
| 5 | Y502H | moderate LOF | LOF | Female | 50 | 0.16 | 32.7 | 2.87 | 4.8 | 96 | 65.1 |
| 6 | N574S | + | LOF | Male | 75 | 0.64 | 35.5 | 3.35 | NA | NA | NA |
| 7 | P613L | moderate LOF | LOF | Female | 75 | 0.87 | 30.9 | 5.19 | 5.5 | 57 | 28.5 |
| 8 | Y775H | moderate LOF | + | Female | 99.6 | 2.4 | 41.8 | 5.01 | 5 | 148 | 162 |
| 9 | P885L | LOF | LOF | Male | 75 | 0.98 | 32.2 | 6.32 | NA | 32.9 | NA |
| 11 | H987R | LOF | + | Female | 75 | 0.83 | 36.2 | 3.39 | NA | 164 | 67 |
| 12 | H987R | LOF | + | Female | 99.6 | 3.6 | 23.3 | 3.76 | 4.9 | 41 | 10.6 |
| 13 | F1154V | + | LOF | Male | 98 | 1.9 | 33.4 | 6.32 | 4.9 | 66 | 10.5 |
| 14 | Y1191N;<br>K1193E | LOF | + | Female | 91 | 1.15 | 34.1 | 3.42 | 7.7 | 60.4 | 63.4 |
| 17 | D1229N | moderate LOF | + | Female | 91 | 1.55 | 31.65 | 2.9 | NA | NA | 26.7 |
| 18 | ΔC1258 | + | LOF | Male | 25 | -0.67 | 79.1 | 4.79 | 5 | 174 | 103 |
| 19 | R1263Q | LOF | + | Male | 91 | 1.43 | 29 | 5.66 | 4.6 | 33.6 | 16.2 |
| 20 | R1263Q | LOF | + | Male | 98 | 2.3 | 28.25 | 5.11 | 5.5 | NA | 50.1 |

## Discussion

Here, we identify ATRNL1 as an integral part of the energy homeostasis control machinery. ATRNL1, a membrane spanning protein which is expressed in MC4R neurons in the PVN, interacts with the MC4Rand potentiates signaling downstream on the receptor in cell culture. We show that the presence of ATRNL1 in MC4R neurons is essential for their activation by melanocortin peptides and required for their normal MC4R-mediated anorexigenic function. We identified multiple variants of ATRNL1 in patients with severe early onset obesity and show that several of those mutations result in impaired MC4R signaling through either Gαs, Kir7.1 or both, possibly contributing to obesity. However, due to the rarity of these variants, additional studies will be needed to establish a causal relationship.

MC4R function is essential to the regulation of food intake and the control of energy homeostasis. As such, the activity of the receptor is tightly regulated through multiple mechanisms including the existence of both endogenous agonists (melanocortins) and inverse agonist (Agouti-related peptide, AgRP) as well as accessory proteins^15,18,20^. Barsh and colleagues previously showed that Attractin, a single transmembrane domain protein, acts as an accessory protein for Agouti, which antagonizes the Melanocortin 1 receptor in the skin to regulate coat color in animals^24^. Disruption of Attractin (encoded by the mahogany locus) in mice blocks the ability of agouti to antagonize the melanocortin 1 receptor in skin, causing hyperpigmentation. Cone and colleagues showed that Attractin-like peptide 1, which is highly homologous to Attractin, interacts with MC4R via its C terminal domain. N- and C-terminal deletion studies revealed that amino acids 1280–1317 of the C-terminus of mouse Atrnl1 interact with mouse MC4R at residues 303–313 in its intracellular C-terminal tail^21^.

In this study we identify ATRNL1 protein as a critical component of the MC4R signaling machinery. We demonstrate that ATRNL1 enhances MC4R signaling both in vitro and in vivo. We show that deletion of ATRNL1 in the PVN or in MC4R neurons results in increased food intake, weight gain and a significant decrease in the anorexigenic effect of MC4R agonists, establishing its physiological relevance. In the absence of ATRNL1, MC4R retains about half of its efficacy in a cell culture system, thus explaining why deletion of ATRNL1 in MC4R neurons does not completely recapitulate the phenotype of *Mc4r* KO mice but rather results in an intermediate phenotype closer to the one observed in heterozygous null mice.

In humans, we identified multiple functional variants of ATRNL present in the GOOS cohort of patients with severe early onset obesity. Genetic association studies in larger cohorts with functional characterization of coding variants, are needed to test whether loss of function variants contribute to variation in BMI in the population and/or the risk of obesity.

We conclude that, ATRNL1 plays an important role in the control of energy homeostasis and may represent a new monogenic cause of obesity.

## Data Availability

All data produced in the present work are contained in the manuscript

## Acknowledgements

This work was supported by a Wellcome Principal Research Fellowship (207462/Z/17/Z), the National Institute for Health and Care Research (NIHR) Cambridge Biomedical Research Centre, the Botnar Fondation, the Bernard Wolfe Health Neuroscience Endowment, the Leducq Foundation grant, and a NIHR Senior Investigator Award (all to I.S.F.). Part of this research has been conducted using the UK Biobank Resource under Application Number 53821 for which analyses were conducted on the Research Analysis Platform. The views expressed are those of the authors and not necessarily those of the NHS, NIHR, or the Department of Health and Social Care.

## Declaration of conflict of interest

I.S.F. has consulted for a number of companies involved in the development of weight loss drugs (Rhythm Pharmaceuticals, Eli Lilly, Sanofi, Astra Zeneca, Nodthera and Novo Nordisk). RDC and University of Michigan hold equity in Courage Therapeutics, and RDC is a founder and board member of the company. All other authors have no competing interests.

## Methods

### Cell culture

MC4Glo cells were a generous gift from Roger Cone’s lab (University of Michigan). HEK293T and MC4RGlo cells were cultured in Dulbecco’s Modified Eagle’s medium DMEM /F-12 supplemented with 2.5% v/v fetal bovine serum, 2.5% v/v calf serum and 1% penicillin–streptomycin solution. Cultures were incubated at 37°C in a humidified atmosphere containing 5% CO2. Cells were transfected using LipoD293 (Signagen) following manufacturer’s recommendations.

### Co-immunoprecipitation

Cells were transfected with 3HA-MC4R and ATRNL1-3Flag. The next day cells were lysed in 0.1% n-Dodecyl-βmaltoside and Protease Inhibitor Cocktail (Sigma catalog #P8340) in PBS at 4°C on a rocking platform for 1 h. Lysates were transferred to a 1.5 ml tubes and centrifuged at 16000 g for 10 min at 4°C. Immunoprecipitation antibody M2 Anti-Flag mouse monoclonal antibody (Sigma #F1804) or anti-HA mouse monoclonal antibody (Biolegend #901501) were added to the supernatant at 1:5000 dilution and incubated overnight at 4°C. Protein-G Dynabeads (Life technologies #10003D) were added to samples and incubated for 1 hour at 4 °C followed by three washes with lysis buffer. Beads were then resuspended in 30 μl LDS sample Buffer with 5% beta-mercaptoethanol and boiled for 5 min.

### Western blot

Proteins were separated by SDS PAGE in 4-12% Bis-Tris gels and transferred on PVDF membrane. Membranes were then blocked with 5% non-fat dry milk in PBST (PBS, 0.1% Tween 20)) for 1 hour at RT, incubated with the indicated antibody at 1:5000 dilution in blocking buffer overnight at 4 °C on a rocking platform, washed 3 times with PBST for 10 min and incubated with HRP-coupled secondary antibody (goat anti-mouse IgG, Biorad #1706516) at 1:5000 for 1 hour at RT. Chemiluminescent substrate was added to membranes and blots were imaged using an Ibright 1500 (Thermo Fisher Scientific).

### ELISA

HEK293T cells were seeded in 24-well plates, and triplicate wells were transfected with 3HA-MC4R/EV or 3HA-MC4R/ATRNL-1-Flag at a ratio of 1-10 respectively. The next day, cells were rinsed with PBS and fixed for 10 min in 4% paraformaldehyde (PFA) at 4°C on a shaker. PFA was washed three times with PBS, and cells were blocked with 5% non-fat dried milk in either PBS (non-permeabilized) or RIPA (permeabilized) for 1 hour at room temperature on a shaker. Cells were incubated with primary antibody anti-HA (mHA.11) diluted at 1/5000 in blocking buffer for 2 hours at room temperature (RT) on a shaker. Cells were washed three times for 5 minutes with PBS RT on a shaker and incubated with secondary antibody [anti-mouse-HRP, 1:5000] in blocking buffer for 1 hour RT. Cells were washed three times for 5 min with PBS before adding 200 µl of 3,3ʹ,5,5ʹtetramethylbenzidine (Sigma-Aldrich). The reaction was stopped with 200 µl of 10% sulfuric acid. 300 L of each sample was transferred to a 96-well plate, and absorbance was measured at 450 nm using a SpectraMax i3 plate reader.

### Stereotaxic surgery

8-10 weeks old ATRNL1^Fl/Fl^ micewere anesthetized with 2.5% isoflurane inhalation before being placed on a stereotaxic apparatus (David Kopf Instruments, Tujunga, CA, USA). After shaving and standard disinfection of the surgical site using iodine, an incision was made to expose the skull, and a hole was drilled. The coordinates to target bilaterally the PVN were measured using the mouse brain atlas from Paxinos and Franklins, A/P – 1 mm, M/L – 0.75mm, D/V – 5mm from skull surface. AAV-GFP or AAV-CRE-GFP were injected bilaterally. Experiments were conducted at least two weeks after surgery to allow recovery and transgene expression.

Following the same procedure previously explained. A 22-gauge stainless steel guide cannula was implanted to the targeted brain regions. For intracerebroventricular (ICV) injections, guide was placed A/P – 0.5mm, M/L – 1.0mm, D/V –1.5mm from skull surface. Canula was inserted in MC4R^Cre^/ATRNL^−/−^, MC4R^Cre^/ATRNL1^Fl/Fl^ or ATRNL1^Fl/Fl^ mice. Guide cannula was fixed in place with dental cement, and mice were allowed to recover for two weeks post-surgery before experiment.

### Feeding studies

10 to 12-week-old mice were used for feeding studies using the BioDAQ food intake monitoring system (Research Diets) for HFD feeding studies and using the fed3 system for CHOW diet studies. Mice were placed in the BioDAQ home cages or single house with fed3 machines for 5 days to allow for acclimation to the new environment and daily handling. After 5 days of acclimation the mice were injected ICV with a saline solution (0.9%) or an agonist (MTII) at 4 μg at nighttime (5pm) or with an antagonist (AGRP) at 4 μg at daytime (9 am). Food intake was continuously measured by the BioDAQ system.

### In situ hybridization

10 to 12-week-old mice MC4^Cre^/ATRNL1^−/−^ were injected bilaterally in PVN with AAV1-CMV-Flex-eGFP following stereotaxic methods previously described. After two weeks recovery, animals were anesthetized with saturated isoflurane in a closed chamber and perfused with 0.1 M phosphate buffer solution (PBS; pH 7.4), followed by 4% paraformaldehyde (PFA) in PBS. After ON in PFA 4% the brain was moved to a sucrose 30% solution until they sunk. After slicing the brain HCR RNA-Fish protocol from Molecular instruments was used to target ATRNL1.

### cFos immunofluorescence

MC4R^CRE^/ATRNL1^−/−^ and MC4R^CRE^/ATRNL1^Fl/Fl^ mice injected IP with saline or 2 mg/kg Setmelanotide. After 2 hours of treatments, animals were anesthetized with saturated isoflurane in a closed chamber and perfused with 0.1 M phosphate buffer solution (PBS; pH 7.4), followed by 4% paraformaldehyde (PFA) in PBS. After ON in PFA 4% the brain was moved to a sucrose 30 % solution until they sunk. Samples were embedded in Tissue-Tek OCT compound (Sakura Finetechical Co., Ltd., Tokyo) and sectioned at 35 µm thickness using a cryostat (Leica Biosystems, Buffalo Grove, IL, USA). Free-floating sections were washed in PBS two times and incubated with blocking buffer containing 5% normal goat serum in PBS with 0.5% Triton X-100 for 1 h followed by rabbit polyclonal anti-cFos antibody (Catalog (Cat.) No. sc 52, Santa Cruz Biotechnology, TX, USA) in blocking solution (1:1000) overnight at 4 °C. After washed with PBS containing 0.1% Tween-20 (PBST), the sections were incubated in 1:500 Alexa Fluor 594 goat anti-rabbit IgG (R37117, Thermo Fisher Scientific) for 1 h at room temperature. The sections were washed in PBST five times and mounted onto superFrost slides (Fisher Scientific, Pittsburgh, PA, USA), air-dried at room temperature in a dark room and coverslipped with Prolong diamond antifade mountant with 4,6-diamidino-2 phenylindole (Molecular Probes by Life Technologies, Carlsbad, CA, USA). For cell counting, three different sections of the ARC of hypothalamus in each mouse were examined using × 20 magnification. cFos-positive cells were manually counted from entire ARC from five mice per group using the Olympus IX3 microscope system and Olympus Cellsens Dimension software.

### cAMP assay

HEK293T or HEK 293T MC4R glo 22F cells were seeded in 6 well plates and transfected with pGloSensor-22F (50%), GFP or ATRNL1-3Flag (50%) and HiBiT-MC4R (5%) for HEK 293T and GFP or ATRNL1-3Flag (100%) for HEK 293T MC4R glo 22F cell lines. Cells were then lifted using TrypLE Express (Thermo Fisher Scientific) and resuspended in 1 ml DMEM/F12 and count for a final concentration 2×10^6^ cells per ml. 10 µl of cell suspension was transferred to wells of a white clear-bottom 384 well plate and left to adhere overnight. The next day, 10 µl of 1.2 mg/ml D-luciferin (GoldBio, CAS 115144-35) in Serum free media DMEM F12 was added to each well and allowed to incubate for a of 2 h. A separate 384 well plate with the agonist alpha MSH in Serum free media DMEM F12 was prepared at varying concentrations and placed in the FLIPR Tetra automated kinetic plate reader (Molecular Devices) together with the plate containing the cells. Luminescence was measured in every well at a sampling rate of 4 s. After 1 min of basal luminescence recording, agonist was injected, and luminescence was measured for an additional 14 min. Each condition was run in five plicates, and experiments were repeated independently at least three times.

### β-arrestin recruitment

HEK cells, seeded in 6 well plates, were transfected with HA-MC4R-LgBiT (5%) and either SmlBiT-b-arrestin2, SmlBiT-β-arrestin1 (5%) with either empty vector or ATRNL-1-3Flag. The next day 20000 HEK Cells were plated in a white 384-well plate (clear bottom), Nanobit substrate was added following Promega recommendations, and the plate was read with a FLIPR Tetra automated kinetic plate reader (Molecular Devices). A separate plate with agonist was prepared and placed in the FLIPR Tetra. Luminescence was measured in every well at a sampling rate of 3 seconds. After 5 minutes of recording to allow measurement of the basal luminescence signal, the agonist was injected, and luminescence was measured for an additional 15 minutes. Baseline signal was subtracted for each well. Alpha MSH was injected at a final concentration of 1 μM for the higher concentration and then a dilution 10 was applied between the different concentration. Each condition was run in triplicate and experiments were repeated independently at least 3 times.

### Automated Patch Clamp Recordings

Automated patch clamp recordings were conducted with precision using the SyncroPatch 384 PE (Nanion Technologies, Munich, Germany), as previously described^35^. The cellular recordings were obtained from an inducible T-REx HEK 293 cell line (Thermo Fisher Scientific), specifically designed to express the melanocortin 4 receptor (MC4R) alongside a carefully engineered mutant form of the inwardly rectifying potassium channel Kir7.1 (M125R). The cell culture environment was controlled and maintained at 37°C in a humidified incubator with a 5% CO_2_ atmosphere. The cells were nurtured in Dulbecco’s Modified Eagle’s Medium (Invitrogen), supplemented with 10% fetal bovine serum (source: Atlanta Biologicals, Flowery Branch, GA) and 1% antibiotic-antimycotic (Thermo Fisher Scientific) to prevent contamination. For the transfection procedure, cells were seeded in sterile 6-well plates (Thermo Fisher Scientific) at a density of 5 × 10^5^ cells per well and were transfected with 1 μg of either an empty vector (control), wild-type (WT), or mutant cDNA of ATRNL1. This transfection was carried out using X-TremeGENE HP DNA Transfection Reagent (Roche Diagnostics), strictly following the manufacturer’s detailed protocol to ensure optimal efficacy. Following a 48-hour incubation period after transfection, the cells underwent a brief treatment with TrypLE (Thermo Fisher Scientific) for 5 minutes to detach them from the plate surface. The cells were then centrifuged at 200g for 5 minutes, resulting in a pellet that was subsequently resuspended in an external recording solution to achieve a final density of 500,000 cells/mL. This suspension was then transferred to the instrument cell hotel, which was carefully maintained at 10°C while being agitated at 200 rpm until the experimental recordings began.

Recordings were performed in a whole-cell voltage clamp configuration using 4-hole, 384-well recording chips, characterized by a medium resistance range of 2–4 MΩ. The external recording solution contained (in mM): 140 NaCl, 4 KCl, 2 CaCl_2_, 1 MgCl_2_, 10 HEPES, and 5 glucose, adjusted to a pH of 7.4 (using NaOH, ∼300 mOsm). Effective cell sealing was achieved with a modified external solution containing 6 mM CaCl_2_, facilitating optimal electrical access. The internal electrode solution comprised (in mM): 110 KF, 10 KCl, 10 NaCl, 10 HEPES, and 10 EGTA, adjusted to a pH of 7.2 (with KOH, ∼280 mOsm). Pulse generation and data collection were systematically carried out using PatchController384 V.3.2.0 software (Nanion Technologies). To accurately assess the MC4R-dependent channel closures or activations induced by AgRP or α-MSH, a hyperpolarization step to -140 mV was applied for 200 ms, starting from a holding potential of -60 mV, and repeated every 5 seconds. During each hyperpolarization step, the peak inward current amplitude of Kir7.1 was carefully measured and normalized against the remaining currents observed after the addition of 10 mM BaCl_2_, which fully blocks the channels, ensuring accurate quantification. Peptides were delivered at various concentrations across the 4-hole, 384-well recording chip. The resulting electrophysiological responses were rigorously analyzed by calculating the ratios of peak currents in the presence (I_compound_) and absence (I_0_) of the test peptides^36^. Additionally, concentration–response profiles were established to accurately determine the respective EC_50_ or IC_50_ values indicative of activating or inhibiting peptide responses. Data analysis and graphical representation were performed using DataController384 V.3.2.0 software (Nanion Technologies) along with GraphPad Prism V.10.5.0, where concentration–response curves were fitted to a three-parameter sigmoidal model for more refined interpretation of the results.

### Human studies

All studies were approved by the Multi-Regional Ethics Committee and the Cambridge Local Research Ethics Committee (MREC 97/21 and REC number 03/103). Each subject (or their parent for those under 16 years) provided written informed consent; minors provided oral consent. All studies were conducted in accordance with the Declaration of Helsinki. People with severe obesity (BMI>3 SD above the mean for age and sex) of early onset (< 10 years) were recruited to the Genetics of Obesity Study (GOOS) (www.goos.org.uk).

### Rare variants from Genetics of Obesity Study

Rare coding variants were interrogated in 1,623 exomes from participants recruited to the Genetics of Obesity Study (GOOS), comprising 927 exomes from the SCOOP cohort of white British participants^29^ and an additional 696 exomes from participants of white British or diverse reported ancestries. After splitting multiallelics and left-normalisation, we annotated variants using Ensembl Variant Effect Predictor (VEP; Ensembl v111, GRCh37 or GRCh38) for gnomAD v2.1 exome allele frequencies in each genetic ancestry group^37^ and predicted consequences with respect to *ATRNL1* transcript ENST00000355044.8 (MANE Select v1.4; coding sequence equivalent to RefSeq transcript NM_207303.4).

### Rare variants from UK Biobank 500K genomes

This research was conducted using the UK Biobank Resource^38^ under application 53821. UK Biobank recruited 500,000 people from across the UK aged between 40-69 years in 2006-2010^38^. We interrogated the *ATRNL1* gene region of the UK Biobank 500K whole-genome sequencing joint variant-calls produced using Illumina DRAGEN v3.7.8 (pVCFs, UKB Field 24310). We obtained variants in *ATRNL1* transcript ENST00000355044.8 exons plus 20bp 3’/5’ exon-flanking regions (chromosome 10, block files b5755-b5797; Field 24310) followed by multiallelic variant splitting and left-normalisation (bcftools v1.11) and variant annotation for predicted consequences in *ATRNL1* transcript ENST00000355044.8 (VEP, Ensembl v111, GRCh38). Body mass index (BMI) was obtained from participants’ first visit to a UK Biobank assessment center (Field 21001, instance 0). To obtain a group of unrelated white British genomes for subset analysis, genetic ethnic grouping was obtained from Field 22006 (self-reported ‘White British’ and tight cluster in genotype principal-component analysis), relatedness was obtained from the UK Biobank utility (ukbgene rel) and one person was excluded from each related pair among ‘white British’ genomes (kinship ≥ 0.0442, KING, third-degree kinship or closer). Our subset of unrelated white British genomes comprised N=155,929 males and N=180,540 females (Field 22001).

The null hypothesis that mean BMI did not differ between carriers and non-carriers was tested using linear regression with adjustment for covariates, glm(phenotype ∼ carrier_indicator + [covariates], family=“gaussian”) with covariates sex, age and age^2^.

