## Supplemental Figure 1 for "Attractin-Like Protein 1 (ATRNL1): An Essential Partner of MC4R to Regulate Body Weight"

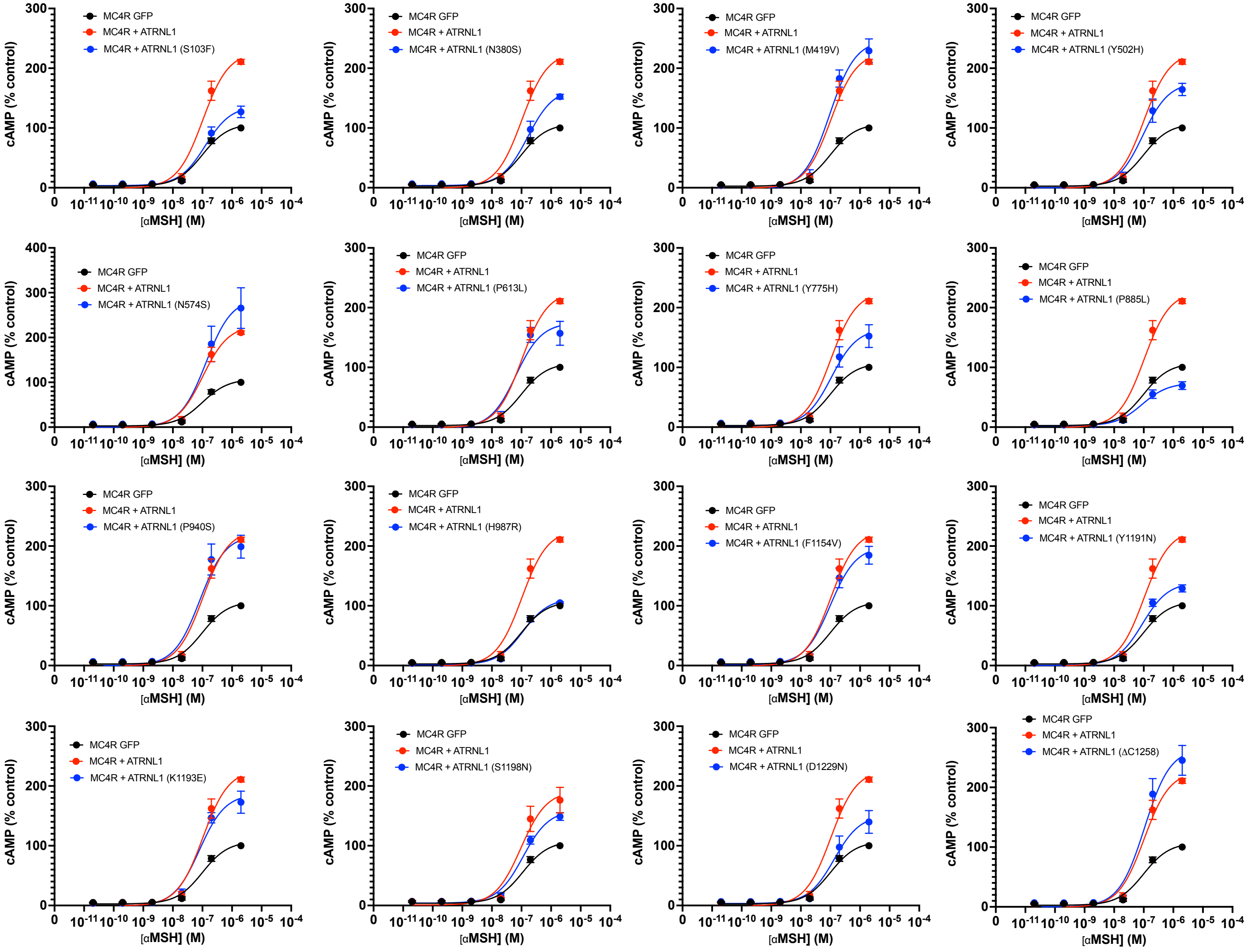


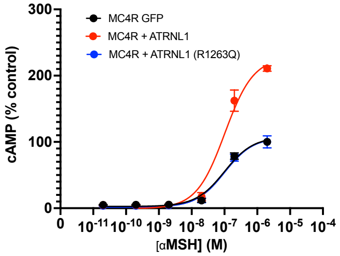


**Figure S1: cAMP responses in cells expressing MC4R with GFP (control), ATRNL1 or indicated ATRNL1 mutant**
