## Supplemental Figure 2 for "Attractin-Like Protein 1 (ATRNL1): An Essential Partner of MC4R to Regulate Body Weight"

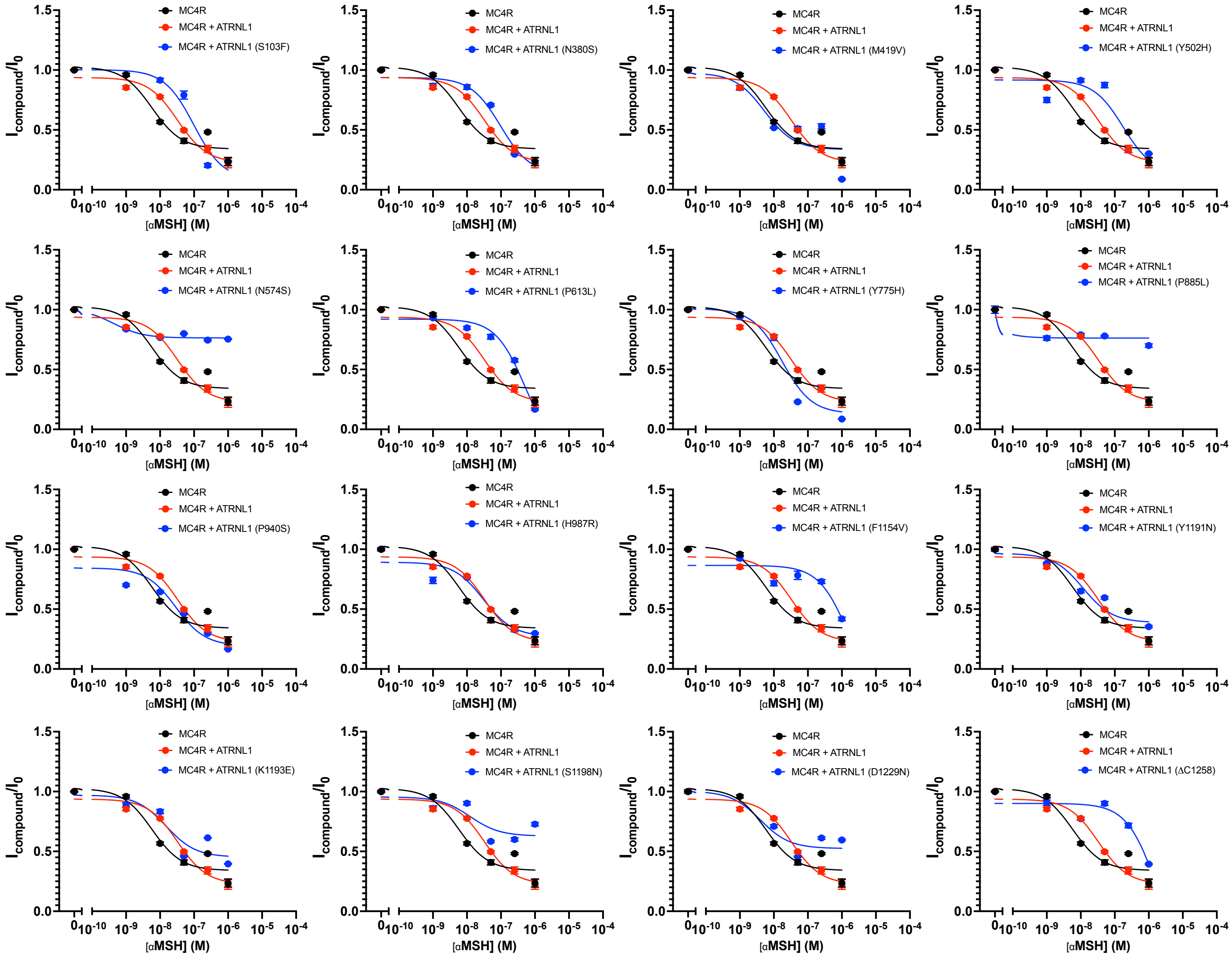

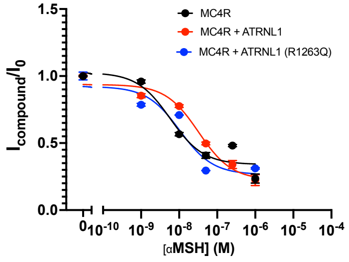


**Figure S2: Potassium currents in response to cAMP stimulation in cells expressing MC4R, Kir7.1 and empty vector, ATRNL1 or indicated ATRNL1 mutant**
